# Covid-19 Epidemiological Factor Analysis: Identifying Principal Factors with Machine Learning

**DOI:** 10.1101/2020.06.01.20119560

**Authors:** Serge Dolgikh

## Abstract

Based on a subset of Covid-19 Wave 1 cases at a time point near TZ+3m (April, 2020), we perform an analysis of the influencing factors for the epidemics impacts with several different statistical methods. The consistent conclusion of the analysis with the available data is that apart from the policy and management quality, being the dominant factor, the most influential factors among the considered were current or recent universal BCG immunization and the prevalence of smoking.

## 1 Introduction

A possible link between the effects of Covid-19 pandemics such as the rate of spread and the severity of cases; and a universal immunization program against tuberculosis with BCG vaccine (UIP, hereinafter) was suggested in [1] and further investigated in [2]. Here we provide a factor analysis based on the available data for the first group of countries that encountered the epidemics (Wave 1) by applying several commonly used factor-ranking methods.

The intent of the work is to repeat similar analysis at several different points in the time series of cases that would allow to make a confident conclusion about the epidemiological and social factors with strong influence on the course of the epidemics.

## 2 Data

### 2.1 Purpose of the Analysis

The zero time of the start of the global Covid-19 pandemics was defined in [2] as:

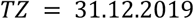

A number of known factors are expected to have strong influence on the course of the epidemics in the national jurisdictions was identified as:

The time of the introduction; demographics; social, tradition, lifestyle; the level of economic and social development; the quality and efficiency of the healthcare system and the quality of policy making and execution.

The purpose of the analysis is to develop and verify the methods that would allow to identify the main factors, different and in addition to the known ones, that have significant influence on the course and the impact of the epidemics based on the available data.

### 2.2 Data

In the first iteration of the analysis we will use only the cases of the first wave that had sufficient time to develop; of those was selected a subset of cases satisfying these conditions:

1. The countries in the set are of a similar development level, thus excluding the influence of the level of prosperity and development; and
2. Certain level of confidence can be expected from the published data.

This selection resulted in the list of 18 cases (Table 1), including one provincial jurisdiction in Canada (Ontario) and a city in the USA (New York City). The time point at which the data was collected was 05.04.2020 i.e. approximately TZ + 3m i.e. approximately 2 months of development for the Wave 1 group of countries.

**Table 1.** **Wave 1 factor analysis data** (recorded: 05.04.2020) **Sources:** World BCG atlas [4] Google coronavirus map [5] World data: Smoking [6] World population data [7] Health Canada Covid-19 updates [8] NYC Covid-19 stats [9] Other [4 – 12]

| *Case* | *Factors* |  |  |  |  | *MV (log m.p.c)* |
| --- | --- | --- | --- | --- | --- | --- |
|  | *Policy* | *UIP* | *Smoking* | *Density* | *Age* |  |
| Taiwan | 0 | 0 | 0.17 | 6.73 | 0.425 | -2.252 |
| Japan | 0.1 | 0 | 0.337 | 3.47 | 0.484 | -0.889 |
| Singapore | 0 | 0 | 0.165 | 83.58 | 0.422 | 0.138 |
| Australia | 0.1 | 0.15 | 0.149 | 0.032 | 0.379 | 0.289 |
| South Korea | 0.1 | 0 | 0.498 | 5.12 | 0.418 | 1.766 |
| Finland | 0.15 | 0.15 | 0.209 | 0.18 | 0.431 | 2.201 |
| Canada | 0.25 | 0.45 | 0.177 | 0.04 | 0.411 | 2.609 |
| Ontario (Canada) | 0.25 | 0.5 | 0.129 | 0.14 | 0.398 | 2.678 |
| Sweden | 0.2 | 0.3 | 0.304 | 2.4 | 0.457 | 3.776 |
| Germany | 0.15 | 0.1 | 0.206 | 0.25 | 0.411 | 5.274 |
| UK | 0.5 | 0.35 | 0.199 | 2.81 | 0.405 | 6.022 |
| France<sup>1</sup> | 0.45 | 0.3 | 0.298 | 1.19 | 0.423 | 6.601 |
| Italy | 0.5 | 0.5 | 0.292 | 2.06 | 0.449 | 7.983 |
| Spain | 0.5 | 0.45 | 0.283 | 0.94 | 0.473 | 7.988 |
| Belgium | 0.35 | 0.5 | 0.16 | 3.76 | 0.413 | 6.814 |
| California | 0.25 | 0.5 | 0.116 | 2.51 | 0.36 | 3.407 |
| New York | 0.5 | 0.5 | 0.125 | 10.19 | 0.358 | 8.034 |
| USA | 0.5 | 0.5 | 0.195 | 0.36 | 0.383 | 4.638 |

The selection eliminated or significantly reduced the influence of several of the known factors mentioned above, namely: the time of arrival, the level of prosperity and development, and to a considerable degree, demographics (although one related factor, the median age was used in the analysis). There was no easy way to eliminate the influence of the policy and quality of execution of epidemics management that in its turn includes a number of factors such as: general epidemics preparedness, effective deployment and management plans, sufficient resources, informed and trained personnel and so on; so it is assumed to be controlled by the policy factor that was assigned manually based on available information. An essential caveat here is that such an assignment would likely itself imply some level of correlation with the observed outcomes, however at the short time of preparation of this analysis it was the best option available. Further studies will be able to produce more objective policy evaluation criteria and methods.

#### Cautions

1. Consistency and reliability of data reported by the national, regional and local health administrations.
2. Alignment in the time of reporting may be an issue due to reporting practices of jurisdictions.

### 2.3 Influencing Factors

In addition to already mentioned policy, the factors considered in this analysis were the following:

**BCG immunization level:** defined in the range 0 – 0.5, with 0 representing band A [3] (i.e. a current universal BCG immunization program) and 0.5 – no UIP (band C). The values in between were assigned in proportion to the time lag between the cessation of UIP and the TZ. Also, some corrections were made for the cases where immunization was administered at an older age or only within a single age cohort i.e. a time span of around 20 years.

Smoking prevalence: in the range 0 – 0.5, defined as the rate of smoking in percent in the population. Where a significant gender difference existed in the population with respect to this factor, the higher value was taken as it’s expected to have a greater influence on the outcome.

**Population density:** the total population / 1 sq.km, divided by 100; we recognize that in some cases such with very large area averaging population over the area may lead to less consistent results; a more detailed analysis with more precisely defined geographic boundaries of the cases is intended for a future study.

**Age demographics:** median age, divided by 100.

### 2.4 Factor Analysis Methods

We used several statistical methods in the factor influence analysis to evaluate the consistency of the obtained conclusions.

Given the large spread within the range of the effect of the epidemics measured as mortality per 1M capita (M.p.c.) in the dataset, a logarithmic scale was used in evaluation of the factor influence, i.e.:

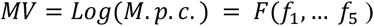

The following methods were used in the analysis:

1. Calculation of correlation between the resulting effect (MV) and the factor
2. Linear regression by one and multiple factors, with evaluation of error
3. Evaluation of feature influence with Random Forest regression [13]
4. Evaluation of feature importance with SelectKBest, a feature ranking method, in sklearn Python library [14].

Method 1 calculates the correlation coefficient between the outcome (MV) and the factor in consideration. An absolute value closer to 1 indicates stronger correlation between the resulting effect and the factor.

Method 2 produces the best fit linear approximation of the resulting effect series with a total deviation (error) from the trend. Comparing the error for different combinations of factors can show which of the factors are most effective in modeling the resulting effect.

Methods 3 and 4 provide a ranking of features with the highest influence on the resulting effect.

## 3 Results

In this section the results are presented for single and multi-factor influence analysis as well as a brief discussion of the obtained results.

### 3.1 Single Factor Analysis

In Table 2 presented correlation and single factor evaluation and ranking results:

**Table 2.** Single factor analysis.

| Factor | Correlation, MV | Linear trend error, MV | RandomForest significance | SelectKbest importance |
| --- | --- | --- | --- | --- |
| Policy | 0.893 | 1.528 | 0.838 | 56.25 |
| BCG UIP | 0.805 | 2.015 | 0.124 | 7.657 |
| Smoking | 0.320 | 3.393 | 0.017 | 0.670 |
| Age demographics | -0.045 | 3.392 | 0.006 | 0.164 |

As can be seen, all methods yielded consistent results with the same rating of the evaluated factors. Apart from the policy factor for which as already discussed, a strong correlation can be expected, the strongest influence factor for this dataset were the BCG immunization, with a strong positive correlation value of *0.81* and the smoking prevalence, *0.32*.

The latter can be expected to be a factor of significance in the epidemics due to already established link with a number of conditions, including respiratory [15]; as a standalone factor it did not show a strong correlation with the outcome, however it can have some influence as a secondary factor as discussed in Section 3.3.

One could have expected a stronger negative correlation for the age demographics, however the result can be explained by a competition of factors of higher susceptibility of the older population group supporting the positive correlation, vs. higher social contact of the younger one stimulating the spread of the infection and thus, acting in the opposite direction and resulting in a lower than expected overall correlation.

### 3.2 Multi-Factor Analysis

In this section we evaluated the effects of the combination of factors with the highest significance: policy, BCG exposure and smoking with the linear regression method. The combination of factors was calculated as a sum of factor values (in the cases with very low population density, a correction was added to account for a slower rate of spread as follows: Canada, Australia: 0.2; Finland, Ontario, USA: 0.1; adding the correction does not change the outcome of the analysis essentially). The results are presented in Table 3.

**Table 3.** Multiple factor analysis: Policy, BCG, Smoking.

| Factor set | Correlation, MV | Linear trend error, MV |
| --- | --- | --- |
| Policy, BCG | 0.867 | 1.226 |
| Policy, Smoking | 0.802 | 1.637 |
| BCG, Smoking | 0.751 | 1.439 |
| Policy, BCG, Smoking | 0.883 | 0.971 |

As can be see seen, the combination of three factors: policy, BCG immunization and smoking has the highest correlation and the lowest linear regression error for the resulting effect. The results also confirm BCG exposure as the second most influential factor among the considered. Indeed, the highest drop in the correlation after removing a factor from the sum is seen for the policy (11.6%) and the lowest, smoking (1.6%). Removing BCG factor results in a reduction of 8.1%.

Figure 1 shows the graphs of the dependency of the resulting effect on the sum of considered factors (Policy + BCG + Smoking): M.p.c., left, a clear exponential trend, and MV = Log(M.p.c), right, a linear trend.

**Fig. 1.**
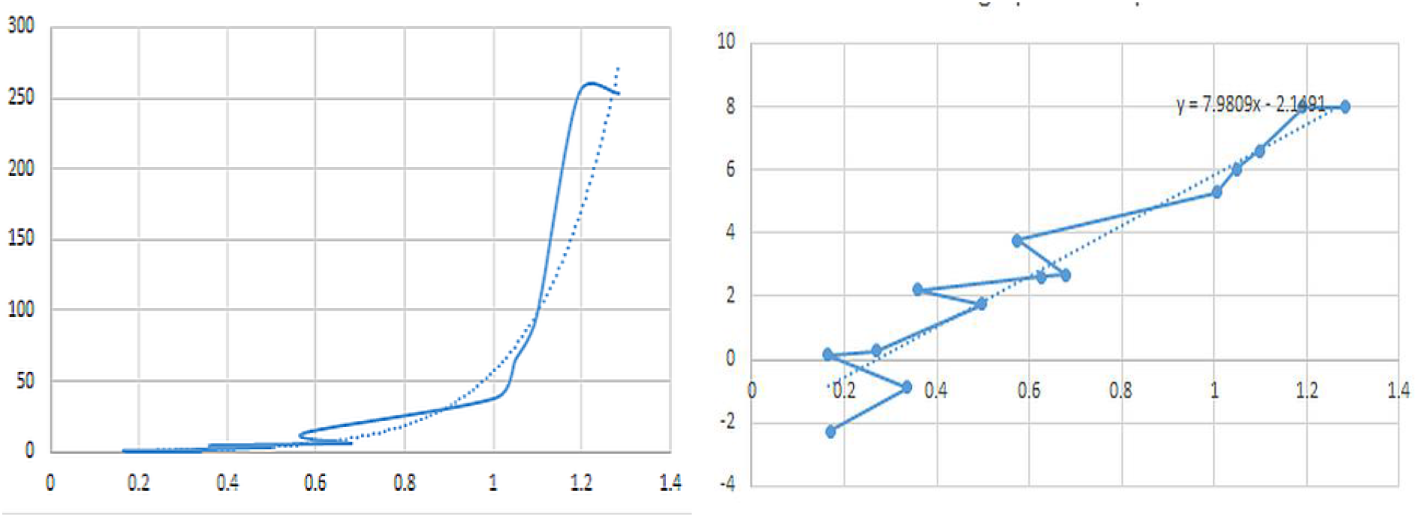
The resulting effect, M.p.c. (left) and MV (right), as function of the factors.

Several outlier cases with a higher than the trend impact clearly seen in the plot on the right can be analyzed in more depth in a future study.

### 3.3 Specific Cases

Interesting observations on the influence of smoking can be derived from a number of specific cases on the role of smoking as a secondary factor in the impact of Covid-19. While may not be sufficient for a statistically confident conclusion, it can provide some directions and rationale for further studies.

#### 1. Asian Group

All countries in this group have similar values of most known factors, including the alignment in time of the onset, the policy and BCG immunization levels (all countries are in group A). The results clearly show that countries with higher smoking prevalence: South Korea and Japan have high impact of the epidemics than those with lower rates (Taiwan, Singapore). Granted, statistical fluctuations are likely in such a data and a confident conclusion can be reached after tracking this trend over a period of time.

#### 2. South America

A similar pattern can be seen with Wave 2 countries in South America. Neighboring countries, with similar known factors such as: Ecuador – Peru, Chile – Argentina but with significantly different smoking rates also show significant difference in Covid-19 impact.

## 4 Conclusion

The analysis of influencing factors and observations obtained on its basis can be useful in understanding the causes impacting the development of the epidemics and possibly developing effective responses and policies on its basis. These early results offer more arguments in support of the hypothesis of some form of BCG immunization protection effect against Covid-19, providing a rationale for further studies of the possible link. We also report the significance of smoking as a secondary factor, consistently confirmed by several methods used in the analysis.

The reported results should not be taken as the definitive statement of a dependence between the investigated factors and the resulting effect, but rather as a demonstration of an approach and methods that over a time would allow to reach a confident conclusion.

## Data Availability

The data referred to in the article is available and included

http://www.bcgatlas.org

https://www.google.com/covid19-map

https://ourworldindata.org/smoking

## Notes

### Competing Interest Statement

The authors have declared no competing interest.

### Funding Statement

This research received no specific funding

### Author Declarations

Revised Common Rule (2018 Requirements) Exempt 4(i)

